# Predicting stress-related disorders from laboratory biomarkers in Finnish electronic health records

**DOI:** 10.1101/2025.09.02.25334838

**Authors:** Anna A. Peltola, Esha Khan, Anna Tirkkonen, Mikaela B. von Bonsdorff, Juulia Jylhävä, Jake Lin

**Affiliations:** Tampere University, Faculty of Medicine and Health Technology, Tampere, Finland; University of Jyväskylä, Faculty of Sport and Health Sciences, Jyväskylä, Finland; University of Jyväskylä, Gerontology Research Center, Jyväskylä, Finland; Folkhälsan Research Center, Helsinki, Finland; Karolinska Institutet, Department of Medical Epidemiology and Biostatistics, Stockholm, Sweden; University of Helsinki, Faculty of Medicine, Helsinki, Finland

## Abstract

**Importance:** Stress-related disorders predict subsequent somatic conditions, suggesting these disorders involve systemic mechanisms, yet evidence from biomarkers remains fragmented across physiological systems.

**Objective:** To investigate whether routinely collected laboratory biomarkers predict the onset of a stress-related disorder and analyze their temporal trends prior to diagnosis.

**Design, setting, and participants:** Nested case-control study using electronic health records from Central Finland (2010-2023). The study included 73,909 participants, aged 34-92 years at baseline, with 6,758 cases of stress-related disorders and 67,151 controls matched to cases on sex and birth year. Average follow-up was 2.8 ± 3.2 years.

**Main outcomes and measures:** Stress-related disorders (International Classification of Diseases, Tenth Revision, category F43 and Z-codes Z73.0 and Z73.3) were the primary outcome. Ten routine laboratory biomarkers were examined: C-reactive protein (CRP), hemoglobin (Hb), glucose, glycated hemoglobin (HbA1c), triglycerides (TG), high-density lipoprotein cholesterol (HDL-C), low-density lipoprotein cholesterol (LDL-C), creatinine (Cr), sodium (Na), and potassium (K).

Temporal trends in these biomarkers were visualized using generalized additive models, and Cox proportional hazards models were used to assess whether biomarker levels measured within one year prior to diagnosis predicted the onset of stress-related disorders.

**Results:** Five biomarkers significantly predicted stress-related disorders within one year prior to diagnosis. Higher potassium (HR 0.73; 95% CI, 0.63–0.84) and higher HDL cholesterol (HR 0.83; 95% CI, 0.74–0.92) levels were associated with reduced risk, whereas higher LDL cholesterol was associated with increased risk (HR 1.10; 95% CI, 1.05–1.16). Higher hemoglobin (HR, 0.98; 95% CI, 0.98–0.98) and higher sodium (HR, 0.96; 95% CI, 0.95–0.98) were also associated with small reductions in risk. No significant associations were observed for remaining biomarkers. According to temporal trend analysis, lipid differences were detectable years before diagnosis, whereas changes in potassium, sodium, and hemoglobin were more transient.

**Conclusions and relevance:** Several routine laboratory markers predict stress-related disorders up to one year in advance, with potassium and lipids showing strongest effects. These findings contribute to understanding the systemic nature of stress-related disorders and point to opportunities for preventive and integrative care approaches.

## Introduction

Stress-related disorders arise in response to stressful or traumatic events.^1^ While defined by psychological criteria, these disorders are predictive of subsequent somatic conditions, including cardiovascular,^2^ immune,^3^ metabolic,^4^ and kidney disease.^5^ These associations suggest that stress-related disorders may involve systemic mechanisms, but evidence from biomarkers remains fragmented across physiological systems.

The physiological stress response is primarily regulated by two neuroendocrine systems: the sympathetic nervous system (SNS) and the hypothalamic-pituitary-adrenal (HPA) axis.^6^ These systems coordinate cardiovascular, immune, and metabolic functions through the release of catecholamines and glucocorticoids.^6–8^ According to the allostatic load model, sustained activation of the SNS and the HPA axis may impair interconnected physiological systems, increasing vulnerability to both psychiatric and somatic conditions.^9–11^

Despite this theoretical understanding, empirical research on stress-related disorders remains limited and uneven across physiological systems. This is particularly true for biomarkers – indicators of biological processes, used to assess disease states, predict progression, and evaluate treatment effects.^12^ Beyond the primary neuroendocrine systems,^13,14^ evidence is strongest for immune markers,^15,16^ promising but less established for metabolic markers,^17,18^ and sparse for other organ systems, such as the renal system.^5^ With few exceptions,^17^ research has rarely addressed pre-diagnostic biomarkers – those that might reflect early physiological changes prior to disorder onset.

Using electronic health records from Central Finland, this study investigates whether routinely collected laboratory biomarkers predict subsequent stress-related disorders. We focus on biomarkers across five categories: immune (C-reactive protein), hematologic (hemoglobin), metabolic (glucose, lipids), renal (creatinine), and electrolyte (sodium, potassium). These biomarkers were selected for their established roles in general health screening and their frequent availability in clinical practice. The selection reflects the broad state of somatic health, allowing for varied connections to stress physiology.

To identify early physiological variations preceding clinical diagnosis, we applied a two-step approach. First, we visualized long-term biomarker trends using generalized additive models (GAMs). Second, we estimated associations between recent biomarker measurements and subsequent diagnoses using Cox proportional hazard models. This approach allowed us to examine whether common clinical biomarkers reflect measurable physiological changes both transiently and years before stress-related disorder diagnosis.

## Methods

### Study population

The electronic healthcare records from the Central Finland Wellbeing Services County include 148,438 individuals (52% female) with laboratory measurements collected between January 1, 2010, and December 30, 2023. During this period, 6,758 (4.6%) received a diagnosis of a stress-related disorder. Among these individuals, the mean age at diagnosis was 56 years and the majority were female.

For this study, we included 73,909 individuals: 6,758 cases of stress-related disorder (9.1%) and 67,151 controls (90.9%). Controls were selected from the cohort to match the joint sex and birth year distribution of the cases and to maintain comparable follow-up periods. All individuals included had at least one laboratory measurement. At baseline, participants were aged 34 to 92 years and were followed for an average of 2.8 ± 3.2 years. Diagnoses and comorbidities were derived from the Finnish version of the *International Classification of Diseases, Tenth Revision* (ICD-10).^1^ The study workflow, depicting participant selection and statistical analyses, is shown in Supplemental Figure 1.

### Study design

We used a nested case-control design within the Central Finland electronic health records.^19^ Controls were frequency matched to cases on sex and birth year by sampling from each sex–birth-year stratum to match the distribution observed in cases.^19^ To ensure comparable follow-up periods, each control was assigned an index date corresponding to a randomly selected diagnosis date from a case within the same stratum.

### Stress-related disorders

We defined stress-related disorders to include acute stress reaction (ICD-10 code F43.0), posttraumatic stress disorder (PTSD; F43.1), adjustment disorders (F43.2), other reactions to severe stress (F43.8), and unspecified reaction to severe stress (F43.9). We also incorporated diagnoses of burnout (Z73.0) and stress not elsewhere classified (Z73.3).

### Laboratory biomarkers

The laboratory biomarkers included C-reactive protein (CRP), hemoglobin (Hb), fasting glucose, glycated hemoglobin (HbA1c), triglycerides (TG), high-density-lipoprotein cholesterol (HDL-C), low-density-lipoprotein cholesterol (LDL-C), creatinine (Cr), sodium (Na), and potassium (K), all measured from blood samples. Extreme values (CRP > 10 mg/L; K > 10 mmol/L) were excluded to minimize the influence of acute comorbidity and technical errors. Laboratory measurements were obtained from the Central Finland Wellbeing Services County, where all analyses are conducted according to standardized protocols of the regional healthcare system.

### Covariates

Participants’ sex, birth year, and somatic comorbid conditions were included as covariates. Comorbidity was defined using the Charlson Comorbidity Index (CCI)^20^ and calculated based on 19 weighted diagnostic codes in the ICD-10.^21^

### Analytical strategy

Our analysis proceeded in two phases: (1) visualizing temporal trends in biomarkers prior to diagnosis and (2) assessing whether biomarker levels predict subsequent stress-related disorder diagnosis.

#### GAM-smoothed temporal trends

We used generalized additive models (GAMs) with smooth spline terms and 95% confidence intervals to visualize biomarker trends before stress-related disorder diagnosis in cases and before index date for controls. This approach identified divergence points between groups and captured nonlinear trends that would guide our predictive modeling strategy.

We initially considered mixed longitudinal models but encountered convergence issues due to sparse repeated measurements per individual, consistent with the known challenges in longitudinal data analysis.^22^

#### Cox proportional hazard models

Building on nonlinear trends observed in the GAMs, we fitted Cox proportional hazards models using the most recent biomarker measurement within a 1-year lookback window.^23^ We first fitted univariable models separately for each biomarker and evaluated their performance using concordance indices from an 80:20 train-test split. As a sensitivity analysis, we also fitted univariable logistic regression models and evaluated them using area under the receiver operating characteristic curve (AUC). Biomarkers that met Bonferroni-corrected significance (p < 0.05 divided by 10 tests) in the univariable models were then included in a multivariable model. All predictive models were adjusted for sex, year of birth, and CCI to account for chronic conditions that may influence biomarker levels.^20,21^

All data were stored and analyzed within the secure server environment of the Helsinki-Uusimaa Hospital District (HUS). Analyses were performed using R version 4.5.0, with packages including *ggplot2 3*.*5*.*2, gridExtra 2*.*3, caret 7*.*0*.*1, survival 3*.*8*.*3*, and *rms 8*.*0*.*0*.

### Ethical considerations

The Human Sciences Ethics Committee of the University of Jyväskylä waived ethical approval for this study. A research permit was obtained from the Central Finland Wellbeing Services County.

According to Finnish legislation (Act on Secondary Use of Health and Social Data 552/2019), ethics committee review was not required as patients were not contacted, and treatment was unaffected. The study was conducted in accordance with the Declaration of Helsinki.

## Results

### Descriptive results

The study included 73,909 participants, of whom 6,758 (9.1%) were cases. At baseline, cases were younger than controls (53.3 ± 12.2 vs. 57.8 ± 12.8 years) and had a higher proportion of females (73.6% vs. 64.4%), despite frequency matching on sex and birth year. These differences are expected: matching balanced the joint distribution of sex and birth year, but proportional sampling within strata can produce moderate deviations in overall sex ratios and mean ages. Baseline age, determined by the first laboratory measurement, was lower in cases, likely due to early onsets stress-related diagnoses. All predictive models were adjusted for sex and birth year to account for these residual differences. Comorbidity levels, as measured by the CCI, were low and comparable between groups. Baseline biomarker levels showed no substantive differences between cases and controls (Table 1; Supplemental table 1 for counts per variable).

**Table 1.** Baseline sample characteristics by case status. Categorical variables are presented as counts (n) and percentages (%); normally distributed continuous variables as mean ± standard deviation (SD); and non-normally distributed variables as median and interquartile range (IQR).

|  | All | Cases | Controls |
| --- | --- | --- | --- |
| **Demographics** |  |  |  |
| Age (mean ± SD) | 57.4 ± 12.8 | 53.3 ± 12.2 | 57.8 ± 12.8 |
| Female, n (%) | 48,207 (65.2%) | 4,975 (73.6%) | 43,232 (64.4%) |
| Male, n (%) | 25,702 (34.8%) | 1,783 (26.4%) | 23,919 (35.6%) |
| **Comorbidity** |  |  |  |
| CCI (median, IQR)<sup>1</sup> | 0.00 (0.00–1.00) | 0.00 (0.00–1.00) | 0.00 (0.00–1.00) |
| **Biomarkers** |  |  |  |
| CRP, mg/L (median, IQR) | 2.00 (1.00–4.00) | 2.00 (1.00–4.00) | 2.00 (1.00–4.00) |
| Hb, g/L (median, IQR) | 138.00 (130.00–147.00) | 137.00 (129.00–146.00) | 139.00 (130.00–147.00) |
| HbA1c, mmol/mol (mean ± SD) | 42.47 ± 11.30 | 41.66 ± 11.39 | 42.56 ± 11.29 |
| Glucose, mmol/L (median, IQR) | 5.70 (5.30–6.20) | 5.60 (5.20–6.10) | 5.70 (5.30–6.20) |
| TG, mmol/L (median, IQR) | 1.10 (0.80–1.60) | 1.10 (0.80–1.60) | 1.10 (0.80–1.60) |
| LDL-C, mmol/L (mean ± SD) | 2.98 ± 0.95 | 3.01 ± 0.92 | 2.97 ± 0.95 |
| HDL-C, mmol/L (mean ± SD) | 1.56 ± 0.47 | 1.56 ± 0.46 | 1.56 ± 0.48 |
| Cr, μmol/L (median, IQR) | 70.00 (61.00–80.00) | 68.00 (59.00–77.00) | 70.00 (61.00–81.00) |
| Sodium, mmol/L (mean ± SD) | 140.79 ± 3.19 | 140.67 ± 2.98 | 140.81 ± 3.21 |
| Potassium, mmol/L (mean ± SD) | 3.97 ± 0.37 | 3.92 ± 0.35 | 3.95 ± 0.37 |

Abbreviations: CCI, Charlson Comorbidity Index; CRP, C-reactive protein; Hb, hemoglobin; HbA1c, glycated hemoglobin; TG, triglycerides; LDL-C, low-density-lipoprotein cholesterol; HDL-C, high-density-lipoprotein cholesterol; Cr, Creatinine.
<sup>1</sup> CCI is reported at the index date instead of at baseline.

### Temporal trends in biomarkers

GAM charts captured temporal trends in biomarker levels throughout follow-up (Figure 1). The visualizations suggested relatively clear separation between cases and controls for certain markers, particularly lipids. Cases tended to have higher triglycerides and LDL-C and lower HDL-C. Other markers showed more variable or nonlinear trends, with abrupt changes or group intersections occurring closer to diagnosis. These patterns informed the selection of a 1-year lookback window for the subsequent Cox proportional hazards models.

**Figure 1.**
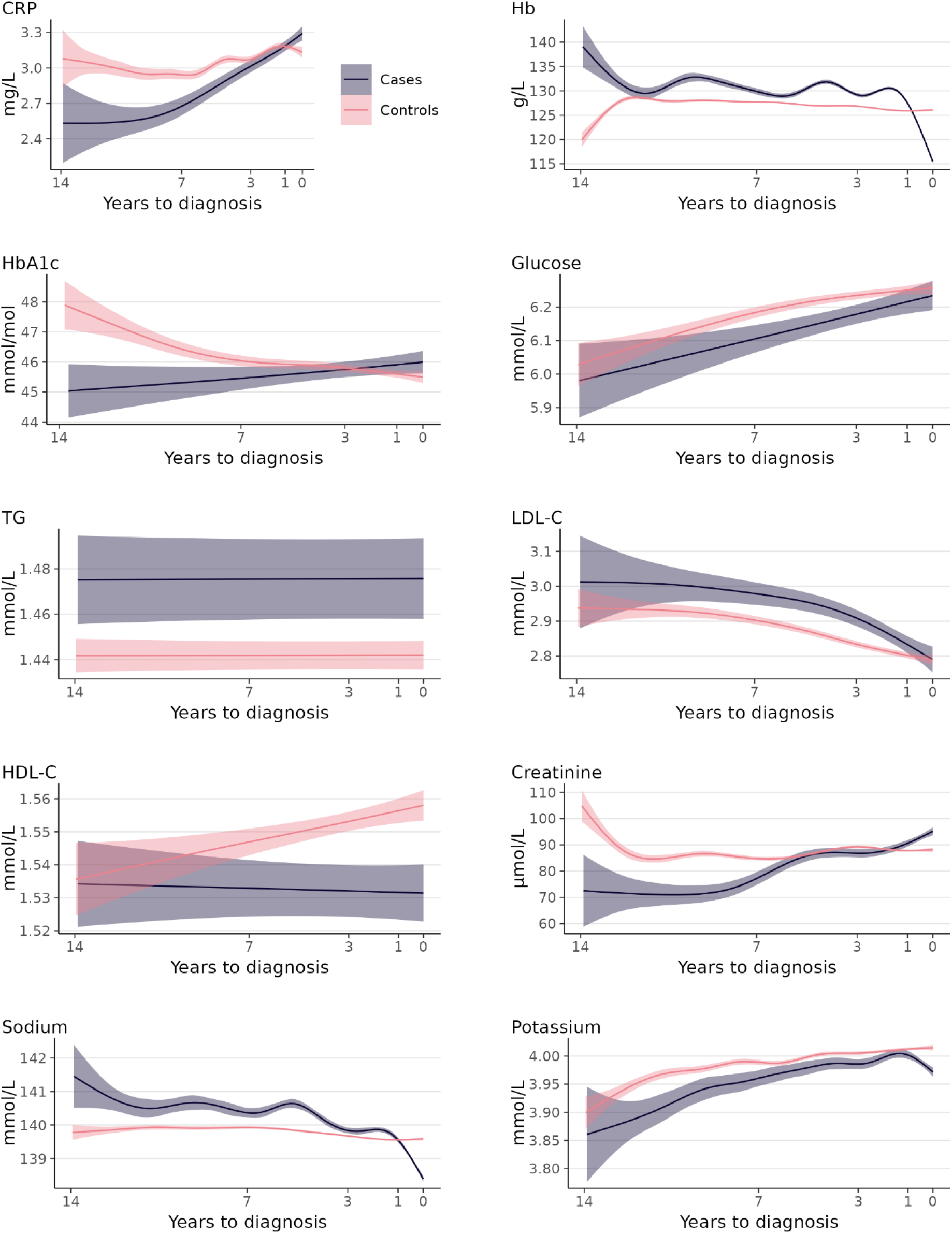
Temporal trends of 10 laboratory biomarkers in the years prior to diagnosis, estimated using generalized additive models (GAMs) with smooth spline terms and 95% confidence intervals. Abbreviations: CRP, C-reactive protein; Hb, hemoglobin; HbA1c, glycated hemoglobin; TG, triglycerides; LDL-C, low-density-lipoprotein cholesterol; HDL-C, high-density-lipoprotein cholesterol.

### Cox proportional hazard models

Cox proportional hazard models were fitted separately for each biomarker (Table 2). Concordance scores from a train-test split indicated stable but limited discriminative performance between cases and controls. Logistic regression analyses with AUC evaluation produced similar results (Supplemental table 2). In univariable analyses, hemoglobin (HR, 0.98; 95% CI, 0.98–0.98), glucose (HR, 0.94; 95% CI, 0.91–0.97), LDL-C (HR, 1.10; 95% CI, 1.06–1.15), HDL-C (HR, 0.86; 95% CI, 0.78–0.94), sodium (HR, 0.98; 95% CI, 0.98–0.99), and potassium (HR, 0.68; 95% CI, 0.63–0.75) reached the Bonferroni-corrected significance threshold (p < 0.05 divided by 10 tests) and were included in a multivariable model (Table 3). All remaining biomarkers were excluded from the multivariable model. In the multivariable model, hemoglobin (HR, 0.98; 95% CI, 0.98– 0.98), LDL-C (HR, 1.10; 95% CI, 1.05–1.16), HDL-C (HR, 0.83; 95% CI, 0.74–0.92), sodium (HR, 0.96; 95% CI, 0.95–0.98), and potassium (HR, 0.73; 95% CI, 0.63–0.84) remained statistically significant (p < 0.05). Higher LDL-C was associated with increased risk of stress-related disorders, whereas higher potassium, HDL-C, hemoglobin, and sodium were associated with reduced risk.

**Table 2.** Results from 10 separate Cox proportional hazards models (Y = stress-related disorder diagnosis), each adjusted for CCI, sex, and birth year. Exposure variables assessed in 1-year lookback window prior to diagnosis.

|  | HR | 95 % CI | p | No. of cases |
| --- | --- | --- | --- | --- |
| CRP | 0.99 | (0.97–0.99) | 0.112 | 2,816 |
| Hb | 0.98 | (0.98–0.98) | <0.001 | 4,892 |
| HbA1c | 1.00 | (1.00–1.00) | 0.156 | 1,955 |
| Glucose | 0.94 | (0.91–0.97) | <0.001 | 2,279 |
| TG | 0.93 | (0.89–0.99) | 0.012 | 2,321 |
| LDL-C | 1.10 | (1.06–1.15) | <0.001 | 2,290 |
| HDL-C | 0.86 | (0.78–0.94) | 0.001 | 2,354 |
| Creatinine | 1.00 | (1.00–1.00) | 0.010 | 4,514 |
| Sodium | 0.98 | (0.98–0.99) | 0.002 | 4,176 |
| Potassium | 0.68 | (0.63–0.75) | <0.001 | 4,186 |

Abbreviations: HR, hazard ratio; CI, confidence interval, p, p-value; CRP, C-reactive protein; Hb, hemoglobin; HbA1c, glycated hemoglobin; TG, triglycerides; LDL-C, low-density-lipoprotein cholesterol; HDL-C, high-density-lipoprotein cholesterol.

**Table 3.** Results from multivariable Cox proportional hazard model^1^ (Y = stress-related disorder diagnosis), adjusted for CCI, sex, and birth year (n = 13,928; no. of cases = 1,576). Exposure variables assessed in 1-year lookback window prior to diagnosis.

|  | HR | 95 % CI | p |
| --- | --- | --- | --- |
| Hb | 0.98 | (0.98–0.98) | <0.001 |
| Glucose | 0.97 | (0.93–1.00) | 0.120 |
| LDL-C | 1.10 | (1.05–1.16) | <0.001 |
| HDL-C | 0.83 | (0.74–0.92) | 0.001 |
| Sodium | 0.96 | (0.95–0.98) | <0.001 |
| Potassium | 0.73 | (0.63–0.84) | <0.001 |

Abbreviations: HR, hazard ratio; CI, confidence interval, p, p-value; Hb, hemoglobin; LDL-C, low-density-lipoprotein cholesterol; HDL-C, high-density-lipoprotein cholesterol.
<sup>1</sup>Analysis was restricted to complete cases for exposure variables.

## Discussion

Several biomarkers predicted stress-related disorder diagnosis within one year prior to onset. Adjusting for somatic comorbidity did not attenuate these associations. Among the electrolytes, higher potassium was associated with a 27% reduced risk, whereas higher sodium showed a smaller reduction. Lipid markers were also predictive: higher HDL cholesterol was associated with moderately reduced risk, while higher LDL cholesterol was associated with a modestly increased risk. Hemoglobin was associated with a small reduction in risk. The remaining biomarkers showed no significant associations. Temporal trend analysis suggested that lipid differences appeared years before diagnosis, whereas changes in potassium, sodium, and hemoglobin emerged closer to diagnosis, suggesting more transient responses to physiological stress.

To our knowledge, our study is the first to have examined electrolyte patterns in stress-related disorders. Higher potassium levels showed a strong protective association, with temporal trend analysis indicating potassium declined close to diagnosis – suggesting responsiveness to acute physiological stress. Two potential pathways could explain these observations. First, acute catecholamine elevations can transiently shift potassium from bloodstream into cells, lowering serum levels.^24,25^ Second, and more plausibly for sustained effects, catecholamines stimulate renin release,^26^ while HPA axis activity stimulates aldosterone secretion.^27,28^ Through the renin-angiotensin-aldosterone system (RAAS), aldosterone induces renal potassium excretion.^29^ The RAAS pathway provides a compelling framework for our potassium findings, especially given that RAAS dysregulation contributes to the same somatic conditions stress-related disorders predict:^29^ cardiovascular,^2^ metabolic,^4^ and renal disease.^5^ In contrast to potassium, the small protective effect of sodium is less consistent with expected physiology, suggesting additional factors may influence its levels.

Our findings contribute to the emerging literature on lipid markers and stress-related disorders. We observed that higher HDL cholesterol reduced the risk for stress-related disorders, whereas higher LDL cholesterol increased the risk, with differences detectable years before diagnosis. These results partially align with the only prior study to predict stress-related disorders from lipid markers,^17^ where our HDL finding was consistent for combined psychiatric disorders but not for stress-related disorders specifically. In contrast, we identified a significant LDL association not observed in that study, underscoring the need for replication. Our results are also consistent with a meta-analysis of PTSD showing higher LDL and lower HDL in patients compared with controls.^18^ The observed associations may reflect glucocorticoid effects on lipid metabolism and HDL’s anti-inflammatory properties.^30,31^ The moderate protective association of HDL and the modest risk increase associated with LDL invite further research.

To our knowledge, hemoglobin has not been previously studied in relation to stress-related disorders. Lower hemoglobin modestly predicted risk, with levels decreasing in the months prior to diagnosis. Given the small effect size, these findings should be interpreted cautiously. Decreased hemoglobin could reflect inflammation-related suppression of red blood cell production.^32^ However, we found no association with CRP, an established marker of low-grade inflammation, which challenges a straightforward inflammatory explanation. Because CRP has been less consistently associated with stress-related disorders than pro-inflammatory cytokines,^15^ the observed hemoglobin changes may reflect immune-related processes not captured by CRP, such as altered iron metabolism,^32,33^ or subclinical somatic conditions not accounted for by our comorbidity adjustments.

These findings indicate that routine laboratory values can predict stress-related disorder diagnosis up to one year in advance, with temporal trend analysis suggesting biological changes occur at different stages before diagnosis. Though causality cannot be determined from observational data, the modest-to-strong effects observed for lipids and potassium have plausible clinical relevance and offer insights into the broad biological underpinnings of stress-related disorders. These findings open research directions for causally informative analysis and determining whether biological patterns could inform preventive and integrative care approaches alongside psychological treatments.

### Limitations

Our study has some limitations. Subclinical conditions may influence laboratory values even after accounting for diagnosed comorbidities. CRP in particular is sensitive to acute inflammation, which can produce transiently elevated values in controls and mask relative sustained changes in cases.

Longitudinal models, which would have allowed individuals to serve as their own controls and addressed this problem, were not feasible due to the limited number of repeated measurements per individual. The definition of stress-related disorder onset is also imprecise, as our follow-up does not fully exclude the possibility of prior diagnosis. While we performed cross-validation, external validation was not possible due to the absence of a suitable independent cohort. Finally, although public healthcare data are broadly representative in Finland, individuals with higher socioeconomic status may be slightly underrepresented.

## Conclusions

Routine laboratory markers predict stress-related disorders up to one year in advance, with potassium and lipids showing strongest effect sizes. These findings advance understanding of the systemic nature of stress-related disorders and suggest opportunities for preventive and integrative care approaches.

## Data Availability

The electronic health records used in this study are not publicly available as they are personally identifying.

## Supplemental material

**Supplemental figure 1.**
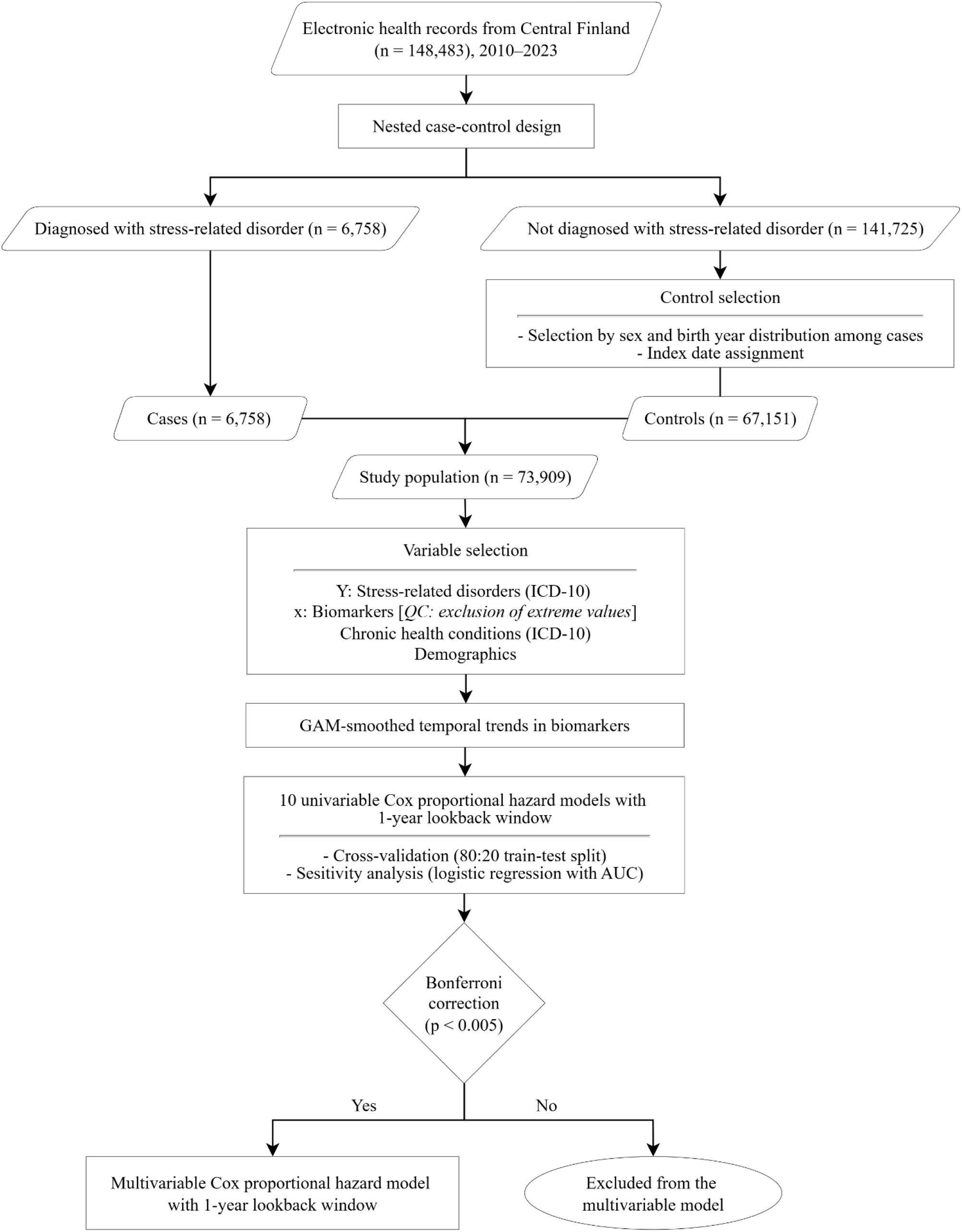
Workflow of the methods section, including study design and statistical analyses. Abbreviations: QC, quality control; GAM, generalized additive model; AUC, area under the receiver operating characteristic curve.

**Supplemental table 1.** Counts of individuals by case status and variable of interest.

|  | All | Cases | Controls |
| --- | --- | --- | --- |
| Age | 73,909 | 6,758 | 67,151 |
| Sex |  |  |  |
| Female | 48,207 | 4,975 | 43,232 |
| Male | 25,702 | 1,783 | 23,919 |
| CCI | 73,909 | 6,758 | 67,151 |
| Biomarkers |  |  |  |
| CRP | 46,744 | 4,883 | 41,861 |
| Hb | 72,442 | 6,664 | 65,778 |
| HbA1c | 33,707 | 3,395 | 30,312 |
| Glucose | 41,454 | 4,334 | 37,120 |
| TG | 42,869 | 4,357 | 38,512 |
| LDL-C | 42,420 | 4,315 | 38,105 |
| HDL-C | 43,170 | 4,377 | 38,739 |
| Creatinine | 66,897 | 6,205 | 60,692 |
| Sodium | 63,225 | 5,960 | 57,265 |
| Potassium | 63,306 | 5,973 | 57,333 |

Abbreviations: CCI, Charlson Comorbidity Index; CRP, C-reactive protein; Hb, hemoglobin; HbA1c, glycated hemoglobin; TG, triglycerides; LDL-C, low-density-lipoprotein cholesterol; HDL-C, high-density-lipoprotein cholesterol.

**Supplemental table 2.** Cross-validation metrics for univariable models based on an 80:20 train-test split.

|  | Concordance |  | AUC |  |
| --- | --- | --- | --- | --- |
|  | *Train* | *Test* | *Train* | *Test* |
| CRP | 0.5482 | 0.5510 | 0.6160 | 0.6028 |
| Hb | 0.6324 | 0.6354 | 0.6535 | 0.6440 |
| HbA1c | 0.5496 | 0.5611 | 0.6340 | 0.6179 |
| Glucose | 0.5814 | 0.5902 | 0.6516 | 0.6303 |
| TG | 0.5721 | 0.5761 | 0.6391 | 0.6158 |
| LDL-C | 0.5849 | 0.5853 | 0.6391 | 0.6132 |
| HDL-C | 0.5938 | 0.5982 | 0.6393 | 0.6133 |
| Creatinine | 0.5661 | 0.5680 | 0.6358 | 0.6207 |
| Sodium | 0.5800 | 0.5855 | 0.6463 | 0.6300 |
| Potassium | 0.5865 | 0.5897 | 0.6406 | 0.6284 |

Abbreviations: AUC, area under the receiver operating characteristic curve.

